# A cautionary note on recall vaccination in ex-COVID-19 subjects

**DOI:** 10.1101/2021.02.01.21250923

**Authors:** Riccardo Levi, Elena Azzolini, Chiara Pozzi, Leonardo Ubaldi, Michele Lagioia, Alberto Mantovani, Maria Rescigno

## Abstract

Currently approved COVID-19 vaccines based on mRNA or adenovirus require a first jab followed by recall immunization. There is no indication as to whether individuals who have recovered from COVID-19 should be vaccinated, and if so, if they should receive one or two vaccine doses. Here, we tested the antibody response developed after the first dose of the mRNA based vaccine encoding the SARS-CoV-2 full-length spike protein (BNT162b2) in 124 healthcare professionals of which 57 had a previous history of COVID-19 (ExCOVID). Post-vaccine antibodies in ExCOVID individuals increase exponentially within 7-15 days after the first dose compared to naïve subjects (*p*<0.0001). We developed a multivariate Linear Regression (LR) model with l2 regularization to predict the IgG response for SARS-COV-2 vaccine. We found that the antibody response of ExCOVID patients depends on the IgG pre-vaccine titer and on the symptoms that they developed during the disorder, with anosmia/dysgeusia and gastrointestinal disorders being the most significantly positively correlated in the LR. Thus, one vaccine dose is sufficient to induce a good antibody response in ExCOVID subjects. This poses caution for ExCOVID subjects to receive a second jab both because they may have a overreaction of the inflammatory response and also in light of the current vaccine shortage.

## Introduction

Currently approved COVID-19 vaccines based on mRNA ^1–3^ or adenovirus ^4^ require a first jab followed by recall immunization. The impact of previous exposure to SARS-CoV-2 on immune response elicited the vaccines has not been assessed.

## Methods

We tested the antibody response developed after the first dose of the mRNA based vaccine encoding the SARS-CoV-2 full-length spike protein (BNT162b2)^1^ in 124 healthcare professionals of which 57 had a previous history of COVID-19 (ExCOVID) (Table 1), as part of an observational study (clinicaltrial.gov NCT04387929) conducted at Istituto Clinico Humanitas in which healthcare professionals were followed for serology and for any occurring COVID-19 related symptoms every three months^5^. We recorded the antibody response to Spike 1/2 with a quantitative test (Liaison SARS-CoV-2 S1/S2 IgG assay (DiaSorin, Italy) which allowed us to evaluate even large amounts of plasma IgG. To predict the IgG response for SARS-COV-2 vaccine, a multivariate Linear Regression (LR) model with l2 regularization (also known as Ridge Regression) was developed. Numerical variables were standardized (z-score algorithm) and the target variable was log transformed due to right asymmetry of the distribution. The subjects without the serological analysis before vaccination were excluded from this analysis (n=11). The final number of subjects analyzed in LR was 113.

**Table 1.** A.B. Demographic distribution of anti-Spike 1/2 IgG plasma levels. C. Days between vaccination and serology post vaccination. *P* values determined using two-tailed unpaired Mann– Whitney test (MW) or two-tailed unpaired Kolmogorov-Smirnov test (KS). NA: not applicable.

| A |  |  | Serology pre VAX |  |  |  |  |  |  |  |
| --- | --- | --- | --- | --- | --- | --- | --- | --- | --- | --- |
|  |  |  | counts | % | min | max | mean | std | TEST | p |
| COVID-19 | NO (Healthy) |  | 67 | 59.29 | 3 | 7.66 | 3.35 | 1.02 | MW | 5E-12 |
|  | YES (ExCOVID) |  | 46 | 40.71 | 3 | 140 | 44.58 | 37.72 |  |  |
| GENDER | F |  | 63 | 55.75 | 3 | 132 | 18.86 | 28.46 | MW | 0.295 |
|  | M |  | 50 | 44.25 | 3 | 140 | 21.74 | 34.99 |  |  |
| Age_class | 21-30 |  | 24 | 21.24 | 3 | 140 | 29.25 | 38.82 | MW | 0.027 |
|  | 31-40 |  | 37 | 32.74 | 3 | 132 | 16.56 | 28.14 | MW | 0.086 |
|  | 41-50 |  | 31 | 27.43 | 3 | 81.7 | 14.76 | 22.91 | MW | 0.074 |
|  | 51-60 |  | 13 | 11.5 | 3 | 123 | 25.73 | 42.37 | KS | 0.146 |
|  | 60+ |  | 8 | 7.08 | 3 | 78.1 | 21.12 | 29.81 | KS | 0.986 |

| B |  |  | Serology post VAX |  |  |  |  |  |  |  |
| --- | --- | --- | --- | --- | --- | --- | --- | --- | --- | --- |
|  |  |  | counts | % | min | max | mean | std | TEST | p |
| COVID-19 | NO (Healthy) |  | 67 | 54.03 | 3 | 104 | 13.29 | 19.73 | MW | 3E-19 |
|  | YES (ExCOVID) |  | 57 | 45.97 | 3 | 4000 | 1055.7 | 1004.2 |  |  |
| GENDER | F |  | 71 | 57.26 | 3 | 3800 | 466.97 | 875.9 | MW | 0.455 |
|  | M |  | 53 | 42.74 | 3 | 4000 | 526.62 | 833.75 |  |  |
| Age_class | 21-30 |  | 30 | 24.19 | 3 | 4000 | 601.13 | 901.65 | MW | 0.015 |
|  | 31-40 |  | 39 | 31.45 | 3 | 3800 | 553.52 | 1046.9 | MW | 0.115 |
|  | 41-50 |  | 33 | 26.61 | 3 | 2320 | 351.12 | 644.59 | MW | 0.234 |
|  | 51-60 |  | 14 | 11.29 | 3 | 2240 | 437.95 | 740.98 | KS | 0.986 |
|  | 60+ |  | 8 | 6.45 | 3 | 1690 | 465.76 | 664.78 | KS | 0.696 |
| CLASS SYMPTOMS<br>(for ExCOVID) | a/paucisymptomatic |  | 15 | 26.32 | 3 | 1800 | 507.73 | 598.39 | KS | 0.029 |
|  | symptomatic |  | 42 | 73.68 | 10.7 | 4000 | 1251.4 | 1051.6 |  |  |
| ExCOVID | Fever | 0 | 30 | 52.63 | 3 | 3800 | 865.65 | 942.76 | MW | 0.034 |
|  |  | 1 | 27 | 47.37 | 10.7 | 4000 | 1266.9 | 1045.2 |  |  |
|  | Low-grade Fever | 0 | 44 | 77.19 | 3 | 4000 | 954.8 | 1060.1 | KS | 0.016 |
|  |  | 1 | 13 | 22.81 | 27 | 2630 | 1397.2 | 717.47 |  |  |
|  | Headache | 0 | 31 | 54.39 | 3 | 2520 | 825.63 | 734.51 | MW | 0.038 |
|  |  | 1 | 26 | 45.61 | 27 | 4000 | 1330 | 1211.4 |  |  |
|  | Cough | 0 | 40 | 70.18 | 3 | 3800 | 921.12 | 959.53 | KS | 0.176 |
|  |  | 1 | 17 | 29.82 | 8.18 | 4000 | 1372.4 | 1064.5 |  |  |
|  | Sore throat | 0 | 31 | 54.39 | 3 | 3330 | 1040.9 | 857.41 | MW | 0.365 |
|  |  | 1 | 26 | 45.61 | 8.18 | 4000 | 1073.4 | 1173 |  |  |
|  | Muscle pain | 0 | 27 | 47.37 | 3 | 4000 | 864.43 | 1100.9 | MW | 0.013 |
|  |  | 1 | 30 | 52.63 | 10.7 | 3330 | 1227.9 | 892.15 |  |  |
|  | Asthenia | 0 | 25 | 43.86 | 3 | 2520 | 717.64 | 784.71 | MW | 0.008 |
|  |  | 1 | 32 | 56.14 | 10.7 | 4000 | 1319.8 | 1086.6 |  |  |
|  | Anosmia/dysgeusia | 0 | 29 | 50.88 | 3 | 4000 | 824.28 | 1044.4 | MW | 0.009 |
|  |  | 1 | 28 | 49.12 | 27 | 3800 | 1295.4 | 918.26 |  |  |
|  | Gastrointestinal disorders | 0 | 33 | 57.89 | 8.18 | 2520 | 902.12 | 753.36 | MW | 0.212 |
|  |  | 1 | 24 | 42.11 | 3 | 4000 | 1266.9 | 1259 |  |  |
|  | Conjunctivitis | 0 | 49 | 85.96 | 3 | 4000 | 1018.2 | 984.56 | KS | 0.896 |
|  |  | 1 | 8 | 14.04 | 27 | 3330 | 1285.8 | 1161.6 |  |  |
|  | Dyspnea | 0 | 41 | 71.93 | 3 | 4000 | 952.52 | 1017.5 | KS | 0.179 |
|  |  | 1 | 16 | 28.07 | 27 | 3330 | 1320.1 | 948.76 |  |  |
|  | Chest pain | 0 | 42 | 73.68 | 3 | 4000 | 989.3 | 988.56 | KS | 0.819 |
|  |  | 1 | 15 | 26.32 | 27 | 3330 | 1241.6 | 1058.9 |  |  |
|  | Tachycardia | 0 | 47 | 82.46 | 3 | 3800 | 982.11 | 870.62 | KS | 0.395 |
|  |  | 1 | 10 | 17.54 | 27 | 4000 | 1401.6 | 1496.3 |  |  |
|  | Pneumonia | 0 | 50 | 87.72 | 3 | 4000 | 1013.9 | 1036.5 | KS | 0.218 |
|  |  | 1 | 7 | 12.28 | 27 | 2240 | 1354.3 | 720.25 |  |  |
| Healthy | Fever | 0 | 63 | 94.03 | 3 | 104 | 13.45 | 20.13 | KS | 0.995 |
|  |  | 1 | 4 | 5.97 | 3 | 30.7 | 10.74 | 13.4 |  |  |
|  | Low-grade Fever | 0 | 62 | 92.54 | 3 | 104 | 14.04 | 20.33 | KS | 0.559 |
|  |  | 1 | 5 | 7.46 | 3 | 6.26 | 3.9 | 1.42 |  |  |
|  | Headache | 0 | 42 | 62.69 | 3 | 104 | 12.68 | 20.73 | MW | 0.438 |
|  |  | 1 | 25 | 37.31 | 3 | 60.5 | 14.3 | 18.28 |  |  |
|  | Cough | 0 | 51 | 76.12 | 3 | 104 | 12.52 | 20.12 | KS | 0.126 |
|  |  | 1 | 16 | 23.88 | 3 | 63.8 | 15.72 | 18.85 |  |  |
|  | Sore throat | 0 | 37 | 55.22 | 3 | 60.5 | 11.02 | 15.69 | MW | 0.256 |
|  |  | 1 | 30 | 44.78 | 3 | 104 | 16.08 | 23.78 |  |  |
|  | Muscle pain | 0 | 58 | 86.57 | 3 | 104 | 12.54 | 19.49 | KS | 0.724 |
|  |  | 1 | 9 | 13.43 | 3 | 55.9 | 18.11 | 21.79 |  |  |
|  | Asthenia | 0 | 56 | 83.58 | 3 | 104 | 12.23 | 19.47 | KS | 0.419 |
|  |  | 1 | 11 | 16.42 | 3 | 49.7 | 18.68 | 21.11 |  |  |
|  | Anosmia/dysgeusia | 0 | 66 | 98.51 | 3 | 104 | 13.4 | 19.86 | KS | 0.687 |
|  |  | 1 | 1 | 1.49 | 6.13 | 6.13 | 6.13 |  |  |  |
|  | Gastrointestinal disorders | 0 | 58 | 86.57 | 3 | 104 | 13.3 | 20.1 | KS | 0.656 |
|  |  | 1 | 9 | 13.43 | 3 | 55.9 | 13.2 | 18.24 |  |  |
|  | Conjunctivitis | 0 | 60 | 89.55 | 3 | 63.8 | 11.5 | 16.12 | KS | 0.419 |
|  |  | 1 | 7 | 10.45 | 3 | 104 | 28.62 | 37.68 |  |  |
|  | Dyspnea | 0 | 64 | 95.52 | 3 | 104 | 13.75 | 20.07 | KS | 0.399 |
|  |  | 1 | 3 | 4.48 | 3 | 4.5 | 3.5 | 0.87 |  |  |
|  | Chest pain | 0 | 63 | 94.03 | 3 | 104 | 13.92 | 20.19 | KS | 0.205 |
|  |  | 1 | 4 | 5.97 | 3 | 4.4 | 3.35 | 0.7 |  |  |
|  | Tachycardia | 0 | 62 | 92.54 | 3 | 104 | 14.07 | 20.32 | KS | 0.195 |
|  |  | 1 | 5 | 7.46 | 3 | 4.57 | 3.61 | 0.84 |  |  |
|  | Pneumonia | 0 | 67 | 100 | 3 | 104 | 13.29 | 19.73 | na | na |
|  |  | 1 | 0 | 0 | 0 | 0 | 0 | 0 |  |  |

|  |  | counts | % | min | max | mean | std | TEST | p |
| --- | --- | --- | --- | --- | --- | --- | --- | --- | --- |
| Days between VAX and serology post VAX | Healthy | 67 | 54.03 | 4.8 | 16.9 | 8.9 | 2.91 | MW | 3E-08 |
|  | ExCOVID | 57 | 45.97 | 4.89 | 20.4 | 13.81 | 4.91 |  |  |

## Results

As shown in Fig.1A-B ExCOVID individuals had a much higher antibody response after the first dose of vaccine than naïve subjects (*p*<0.0001), regardless of when they developed the COVID-19. They displayed an exponential increase of anti-Spike 1/2 antibody response within 7-15 days after the first dose of vaccine. The pre-vaccine antibody amount of the ExCOVID population was on average 44.+/-37.7 while that after the vaccine was 1055.7+/-1004.2 (*p*<0.0001) (Table 1), with higher levels in symptomatic ExCOVID (Fig. 1C, *p*=0.028).

**Figure 1:**
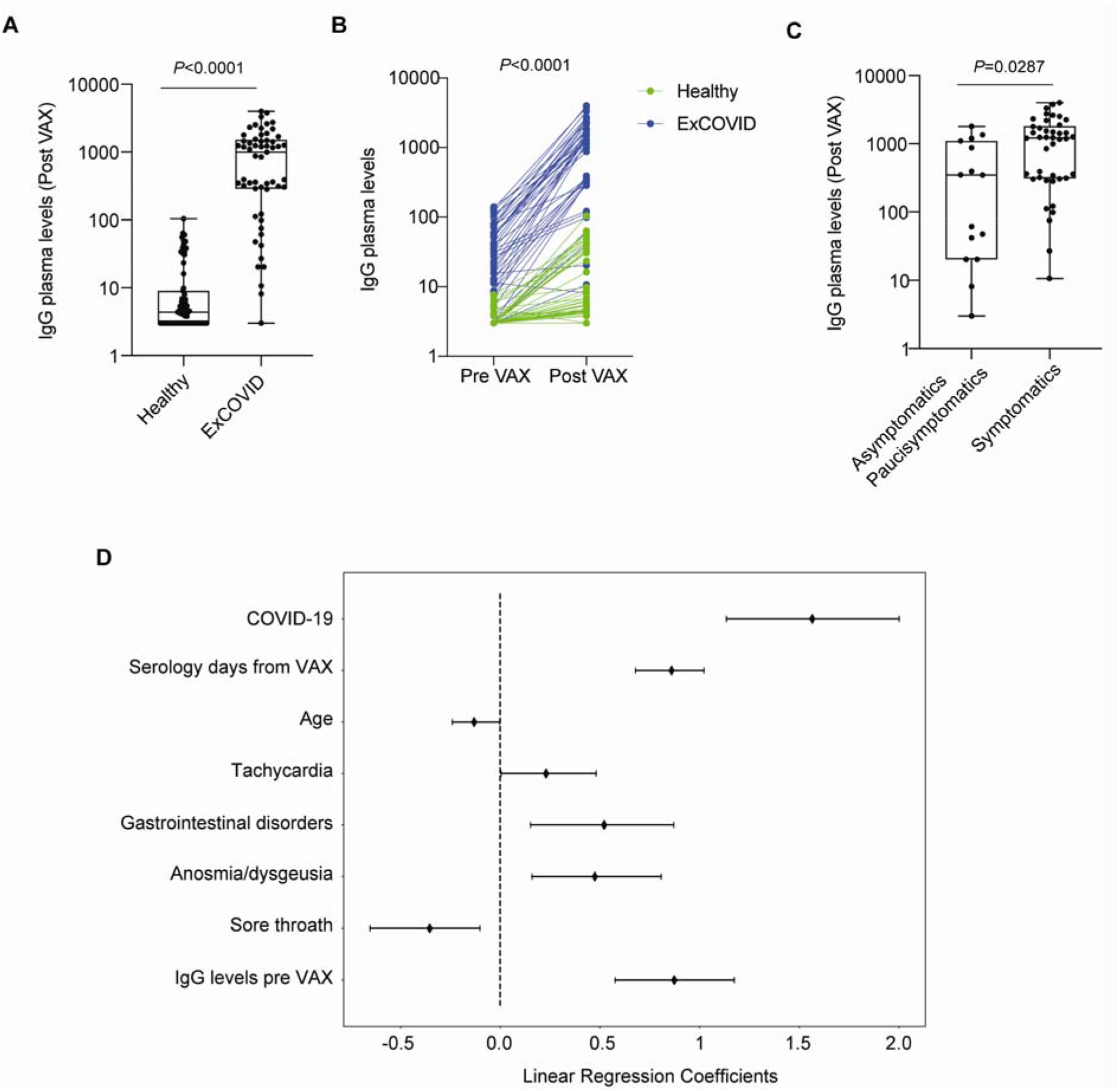
ExCOVID subjects increase exponentially anti-Spike 1/2 IgG levels after the first dose of vaccine. A, anti-Spike 1/2 IgG plasma levels in healthy (n=67) and ExCOVID individuals (n=57) measured after the first dose of vaccine. B, anti-Spike 1/2 IgG plasma levels in healthy (n=67) and ExCOVID individuals (n=46) measured before and after the first dose of vaccine. C, anti-Spike 1/2 IgG plasma levels in asymptomatic / paucisymptomatic (n=15) and in symptomatic (n=42) ExCOVID individuals measured after the first dose of vaccine. D, Multivariate linear regression coefficients for the most significant variables (*p*<0.05). Dot points represent the mean values and the lines the 95% CI. The box plots (A, C) show the interquartile range, the horizontal lines show the median values and the whiskers indicate the minimum-to-maximum range. *P* values were determined using two-tailed unpaired Mann–Whitney test (A) or two-tailed Wilcoxon matched-pairs signed rank test (B) or two-tailed unpaired Kolmogorov-Smirnov test (C).

We investigated the relationship between the amount of IgG after vaccination with COVID-19, sex, age and symptoms related to disease. The final LR shows a good prediction of the target variable (R^2^=0.88, F-statistic = 39.18, *p*-value<0.001) and the most significant features were history of COVID-19 (1.48, 95% CI 1.07-1.93), the value of IgG before vaccination (0.87, 95% CI 0.59-1.13), the difference between the date of vaccination and the date of serology post-vax (0.87, 95% CI 0.65-1.03), and age (−0.13, 95% CI −0.24-−0.001) as well as COVID-19 related symptoms: gastrointestinal disorders (0.59, 95% CI 0.16-0.97), anosmia/dysgeusia (0.50, 95% CI 0.14-0.87), tachycardia (0.26, 95% CI 0.02-0.60) and sore throat (−0.35, 95% CI −0.53-−0.11) (Fig. 1D, Suppl. Table1).

## Discussion

The antibody response of ExCOVID patients depends on the IgG pre-vaccine titer and on the symptoms that they developed during the disorder, with anosmia/dysgeusia and gastrointestinal disorders being the most significantly positively correlated in the LR, while sore throat was negatively correlated because 45% non-COVID individuals reported it. Young subjects had a higher antibody response. We previously observed that anosmia/dysgeusia was associated with an increase of antibodies over time, independently of vaccination (Levi et al. submitted). Thus, one vaccine dose is sufficient to induce a good antibody response in ExCOVID subjects and poses caution for a second dose: over stimulation with high amount of antigens could switch-off the immune response due to antigen exhaustion, which occurs in response to several viruses (reviewed in^6^). Alternatively, overactivation of the immune response may drive the development of low-affinity antibodies for SARS-CoV-2 which may foster an antibody dependent enhancement (ADE) reaction when re-exposed to the virus (reviewed in^7^). These results question whether a second shot in ExCOVID subjects is indeed required and suggest to post-pone it while monitoring antibody response longevity. At a time of vaccine scarcity, these findings may have public health implications.

## Data Availability

Data supporting the findings of this study are available within the paper and its Supplementary Information files. All other data are available from the corresponding author upon reasonable requests.

