## Supplementary Table 1 for "A cautionary note on recall vaccination in ex-COVID-19 subjects"

**Supplementary Table 1:** Multivariate linear regression coefficients and exponentiated coefficients for all the variables analyzed.

| | Coeff | 5% | 95% | Exponentiated Coefficients | 5% | 95% | |
|---|---|---|---|---|---|---|---|
| Constant | 2.98 | 2.78 | 3.25 | 19.65 | 16.05 | 25.67 | * |
| IgG levels pre VAX | 0.87 | 0.59 | 1.13 | 2.38 | 1.80 | 3.10 | * |
| Fever | 0.16 | -0.28 | 0.60 | 1.18 | 0.76 | 1.82 | |
| Low-grade Fever | -0.06 | -0.45 | 0.22 | 0.94 | 0.64 | 1.24 | |
| Headache | -0.03 | -0.35 | 0.28 | 0.97 | 0.70 | 1.32 | |
| Cough | 0.04 | -0.25 | 0.34 | 1.04 | 0.78 | 1.41 | |
| Sore throath | -0.35 | -0.53 | -0.11 | 0.70 | 0.59 | 0.89 | * |
| Muscle pain | 0.26 | -0.08 | 0.49 | 1.30 | 0.92 | 1.63 | |
| Asthenia | 0.18 | -0.18 | 0.48 | 1.20 | 0.83 | 1.61 | |
| Anosmia/dysgeusia | 0.50 | 0.14 | 0.87 | 1.65 | 1.16 | 2.38 | * |
| Gastrointestinal disorders | 0.59 | 0.16 | 0.97 | 1.81 | 1.17 | 2.63 | * |
| Conjunctivitis | -0.18 | -0.48 | 0.17 | 0.84 | 0.62 | 1.19 | |
| Dyspnea | 0.17 | -0.10 | 0.48 | 1.19 | 0.90 | 1.61 | |
| Chest pain | 0.11 | -0.14 | 0.38 | 1.12 | 0.87 | 1.46 | |
| Tachycardia | 0.26 | 0.02 | 0.60 | 1.30 | 1.02 | 1.83 | * |
| Pneumonia | -0.11 | -0.48 | 0.23 | 0.90 | 0.62 | 1.26 | |
| Age | -0.13 | -0.24 | -0.001 | 0.88 | 0.79 | 1.00 | * |
| Sex | -0.01 | -0.16 | 0.19 | 0.99 | 0.85 | 1.21 | |
| Serology days from VAX | 0.87 | 0.65 | 1.03 | 2.39 | 1.91 | 2.80 | * |
| Serology days before VAX | -0.003 | -0.01 | 0.01 | 1.00 | 0.99 | 1.01 | |
| COVID-19 | 1.48 | 1.07 | 1.93 | 4.39 | 2.92 | 6.92 | * |

** p<0.05*
